# National age and co-residence patterns shape covid-19 vulnerability

**DOI:** 10.1101/2020.05.13.20100289

**Authors:** Albert Esteve, Iñaki Permanyer, Diederik Boertien, James W. Vaupel

## Abstract

Based on harmonized census data from 81 countries, we estimate how age and co-residence patterns shape the vulnerability of countries’ populations to outbreaks of covid-19. We estimate variation in deaths arising due to a simulated random infection of 10% of the population living in private households and subsequent within-household transmission of the virus. The age-structures of European and North American countries increase their vulnerability to covid-related deaths in general. The co-residence patterns of elderly persons in Africa and parts of Asia increase these countries’ vulnerability to deaths induced by within-household transmission of covid-19. Southern European countries, which have aged populations and relatively high levels of intergenerational co-residence are, all else equal, the most vulnerable to outbreaks of covid-19. In a second step, we estimate to what extent avoiding primary infections for specific age-groups would prevent subsequent deaths due to within-household transmission of the virus. Preventing primary infections among the elderly is the most effective in countries with small households and little intergenerational co-residence such as France, whereas confining younger age groups can have a greater impact in countries with large and inter-generational households such as Bangladesh.

## Introduction

The covid-19 pandemic currently confronts nearly all of the world’s countries. A growing number of governments are enforcing or recommending home quarantines to contain the spread of the virus. As the virus can be transmitted outside and within households, the effects of such measures will depend on the number of transmissions that take place outside and within the household. Evidence shows that the risk of severe disease and mortality increases sharply with age (1, 2). Therefore, the age structure of the population—what proportion are young or old—and the structure of co-residency—how big are households and how old are their members—are two key factors that determine the vulnerability of countries to outbreaks of covid-19, and how effective general and age-specific household confinement policies can be in reducing mortality after an outbreak (3).

## Results

Figure 1 provides estimates of the number of deaths from covid-19 per 100,000 individuals if countries were to experience an outbreak of covid-19 of equal size, more specifically, a random infection of 10% of the population living in private households. Results are shown for 81 countries, covering all regions of the world and are solely based on census-based micro-data on age and co-residence patterns combined with age-specific infect fatality ratios (2). The left-hand segment of each bar provides an estimate of direct mortality of individuals who catch the disease in a 10% random infection of the population (primary infections). The right-hand segment of the bars shows the additional deaths that would occur if all other members of the household become infected too (secondary infections). Lower rates of household transmission would reduce this number of indirect deaths proportionally. The direct effect depends on the age structure of the population; the indirect effect hinges on the size and age structure of households. Combined, they show how, all else equal, national age and co-residence patterns alter the vulnerability of a country to covid-19 outbreaks.

**Figure 1.**
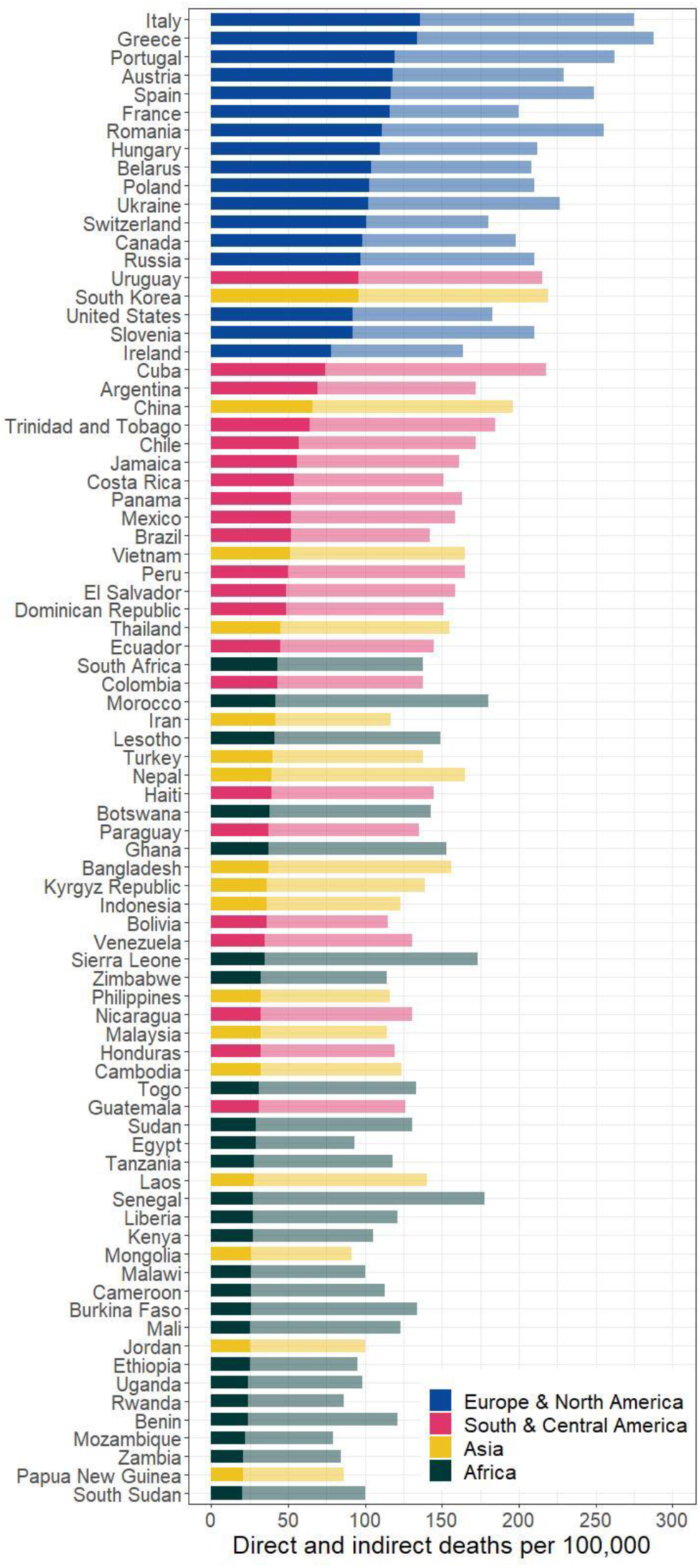
Covid-19 vulnerability to national age and co-residence patterns. Estimated number of direct (dark) and indirect (light) deaths per 100,000 individuals if 10% of the population living in private households were to be infected by covid-19 at random.

The expected direct death rates per 100,000 individuals range from 20 in South Sudan to 136 in Italy. Together with Italy, three Southern European countries—Greece, Portugal and Spain—rank among the top five, followed by the rest of Europe and North America. Latin American countries form a homogenous cluster lower than the European and North American cluster. Asian countries spread all over the range with estimates as high as 96 in South Korea and as low as 25 in Jordan. African countries tend to experience the lowest direct death rates. Where the elderly comprise a large portion of the population, the direct effect is high, whereas direct deaths are much lower where the elderly are vastly outnumbered by younger people.

Mortality due to intra-household contagion (right-hand segment of a bar) does not follow the same order because co-residence patterns differ widely across countries, even among those countries with similar age structures (4, 5, 6, 7). The ratio between indirect and direct effects is a simple indicator of the importance of co-residence patterns, in particular of the elderly, the most vulnerable group. For European and North American countries, direct and indirect deaths are roughly equal. In Latin America, indirect deaths could approximately double the number of direct deaths. The ratio between potential indirect and direct deaths in Asia ranges from 1.3 (South Korea) to 3.2 (Bangladesh). In Africa, indirect deaths would be three to four times the number of direct deaths. Such variation is closely associated with cross-national variation in co-residence patterns and, more specifically, with the number and age of the persons with whom elderly people reside.

Despite differences in ratios, the combined death rate (direct plus indirect) reproduces a broad regional pattern similar to the one observed in direct mortality but with variation in the specific position of countries. For instance, countries with similar direct death rates, such as France and Spain, show remarkably different indirect rates due to higher levels of intergenerational co-residence in Spain. Countries with similar indirect death rates, such as Italy and China, have quite distinct direct death rates, due to differences in their age structure.

### Preventing Primary Infection of Specific Age Groups

Debates exist about the role that specific age-groups, and particularly children, play in the transmission of the virus (8). In addition, countries have adopted age-specific policies such as school closures and extended confinement of the elderly in their homes. This provokes the question to what extent preventing primary infection of certain age groups would reduce the number of deaths that can arise due to within-household transmission of the virus.

The bars in Figure 2 provide information about the expected mortality from direct and indirect deaths if primary infections for specific age groups could be averted. Results are shown for 10 countries chosen to illustrate diverse interactions between age structures and co-residence patterns (see Figure S3 for all countries). The first column shows the total number of deaths per 100,000 population if no age group is excluded from primary infection. The other columns indicate the total number of direct and indirect deaths per 100,000 population if primary infections could be averted for a particular age group.

**Figure 2.**
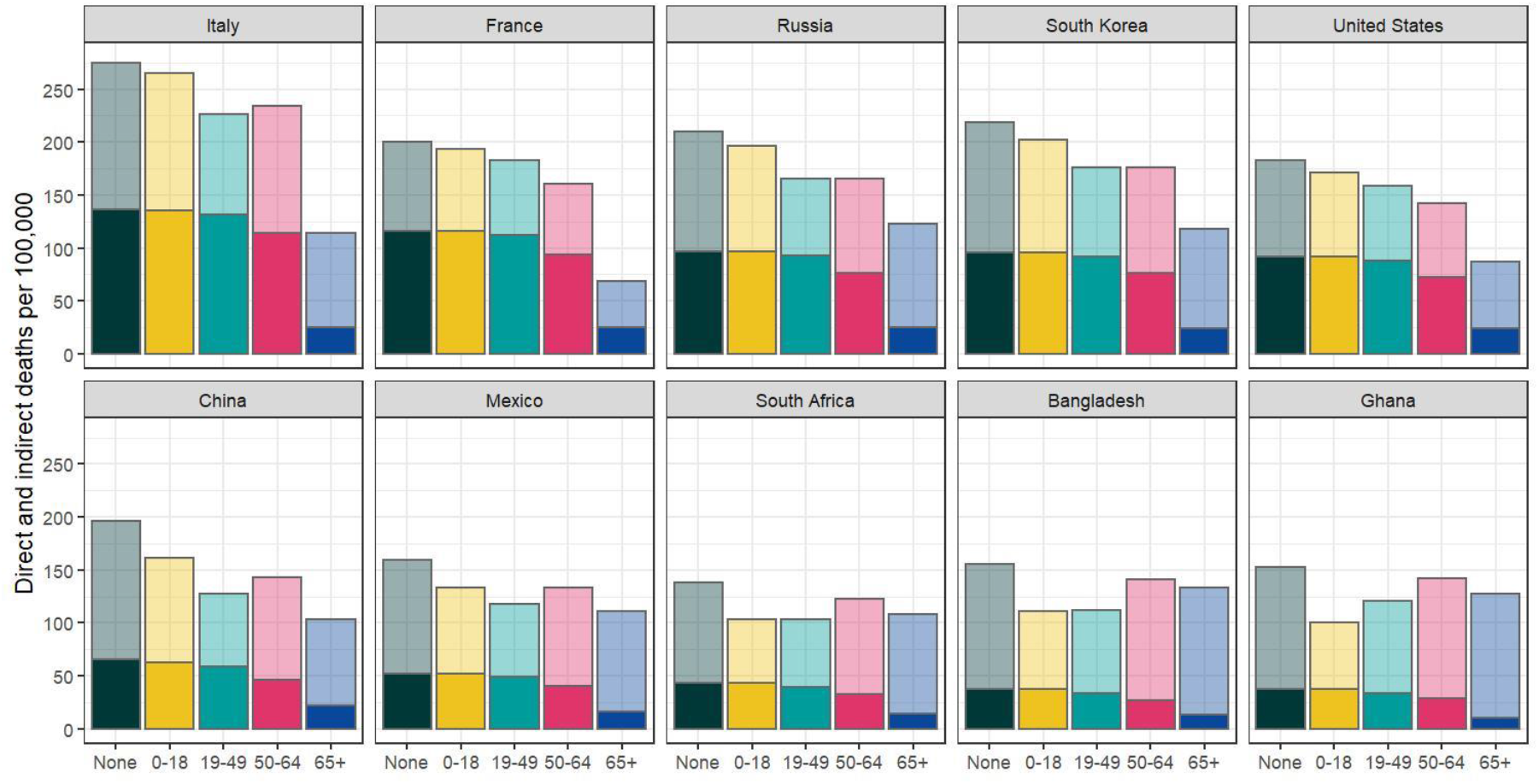
Estimated number of direct (dark) and indirect (light) deaths per 100,000 individuals if primary infections of specific age groups are avoided. Individuals from each age group who were selected in the 10% random draw are recoded as not infected before calculating direct deaths and simulating within household transmission.

Deaths due to primary infection are reduced most in all countries if infections of individuals aged 65 or more are prevented. There is much more variation in how much avoiding primary infections of specific age-groups affects the number of indirect deaths that arise due to secondary infection (within household transmission). In France, indirect deaths are simulated to go down considerably if no person aged 65 or older were to be infected directly. This indicates that elderly persons primarily live with other persons aged 65 or older in France (Figure S4, Figure S5). These co-residence patterns also imply that avoiding primary infections for other age groups have relatively little effect on deaths that emerge due to within household transmission of the virus in France. At the other extreme, there are countries such as Bangladesh where preventing direct infections of the elderly would barely reduce indirect deaths and where avoiding the primary infection of children or adults aged 19–49 has a larger impact on indirect deaths. This result is explained by the high levels of intergenerational co-residence of the elderly together with the fact that children comprise a large share of the total population in these countries (Figure S4, Figure S5). Other countries fall between these two extremes with the United States being similar to France, and Ghana and South Africa resembling Bangladesh. Some cases combine elements from both extremes, such as Italy where both confining the elderly and individuals aged 19–49 reduce indirect deaths. However, in none of the scenarios do indirect deaths go down considerably. This illustrates the double challenge that countries such as Greece, Italy, Portugal and Spain face: the combination of an aged population with inter-generational residence leads to high estimated death rates due to covid-19 but also makes preventing deaths due to within-household transmission of the virus particularly challenging.

## Discussion

In confronting covid-19, epidemiologists should analyze and policymakers should consider how age structure and co-residence patterns in their countries can shape the number of infections and deaths. Differences in age structures put countries at different risk; a less considered factor, co-residence patterns, modulates this risk. In our simulations, which can be considered baseline scenarios before accounting for specific national policies, the proportion dying per 100,000 population is 275 in Italy and 80 in Mozambique (Figure 1). In contrast, if 10% of the population is infected at random and their household members become infected too, the proportion of the population that becomes infected is 28% in Italy and 44% in Mozambique (Figure S2). Because of different age and co-residence patterns, Italy is confronted with more deaths per capita than Mozambique but fewer infections. The effectiveness of policies in one country compared with another country should be evaluated in light of different baseline vulnerabilities.

In countries where the elderly form a large part of the population and primarily live with their generational peers, avoiding the primary infection of elderly people will considerably reduce direct deaths and will also prevent indirect deaths due to within household transmission of the virus. In countries where the elderly form a small part of the population but live together with young people, indirect deaths through infection within households can outnumber direct deaths. Therefore, avoiding primary infections of the elderly will be less effective in reducing deaths because the elderly might still get infected by younger household members. In such cases, measures that reduce or avoid within household transmission of the virus to the elderly become relatively more important to reduce mortality due to covid-19.

## Materials and Methods

Data on age structure and co-residence come from high-quality harmonized census data from IPUMS (9) on individuals living in private households (i.e. individuals living in collective dwellings such as old-age homes are excluded). Mortality is determined by age-specific covid-19 death rates (2). [See supplementary materials (SM) and figs. S7–11 for more details on methods, data, and other robustness checks].

## Data Availability

Data comes from scientific use files of census microdata available at IPUMS international (www.ipums.org)

## Acknowledgments

We thank A. Turu for help with the harmonization and graphic representation of the data. This analysis was supported by the following grants: ERC-2014-StG-637768, RTI2018–096730-B-I00.

## Author Contributions

A.E. formulated research idea; A.E., I.P., D.B. and J.V.W. designed research; D.B. Analyzed data; A.E., I.P., D.B. and J.V.W. wrote the paper.

## Supplementary Information

We provide the following supplementary information I that the editors might consider useful to provide to reviewers. If published, this material (which includes replication code) will be made available on the authors’ own website.

**Figure S1.**
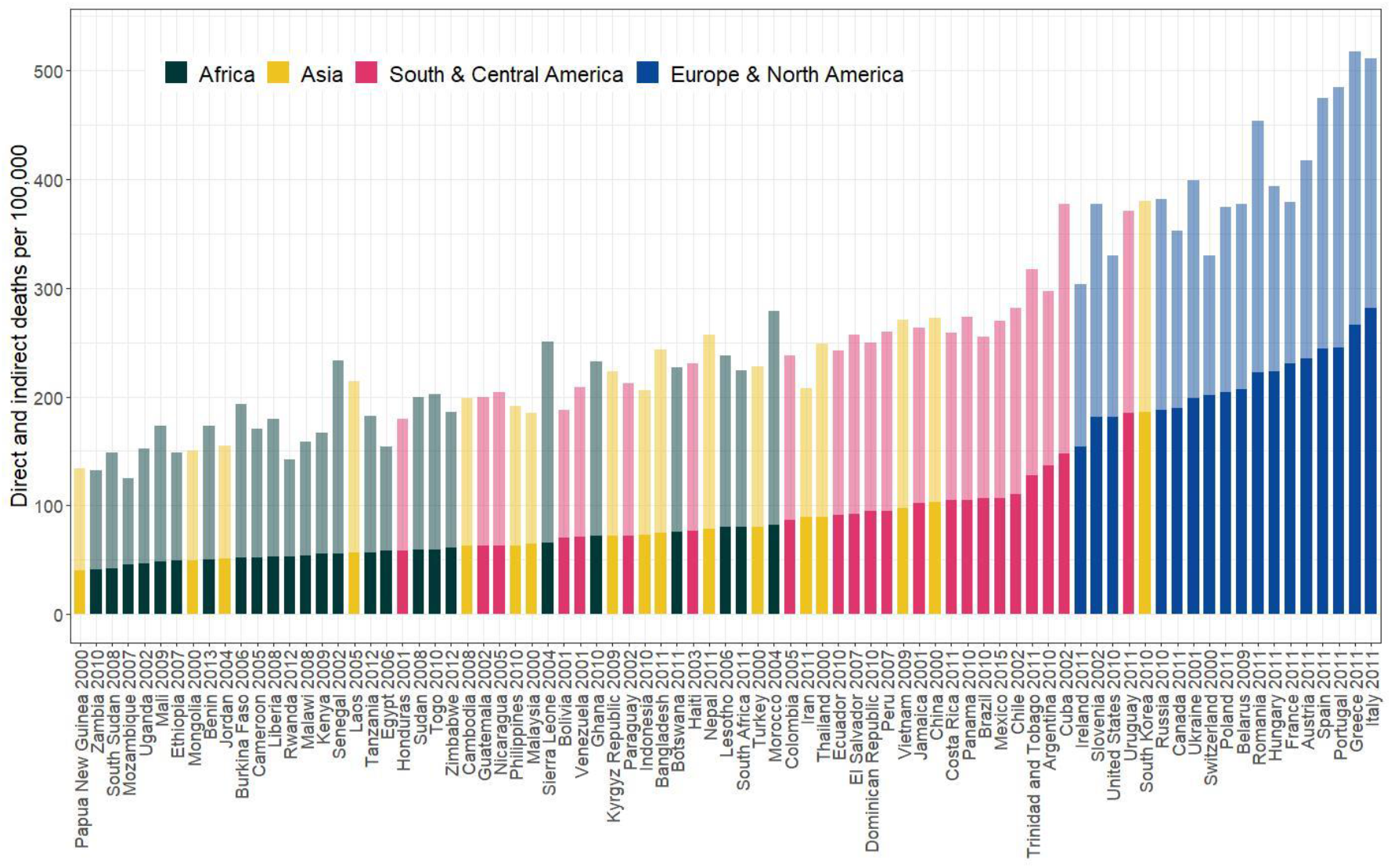
Analysis with 20% primary infections. In our main analysis, we randomly simulated a primary infection rate of 10%. In robustness checks, we simulated a 20% infection rate. Figure S1 displays results from this robustness check. It can be observed that countries with large households move down the distribution of death rates. This is because in several African countries infecting 10% of the population at random already leads to the majority of the population being infected if all household members get infected too. Simulating additional primary infections therefore leads to relatively fewer additional secondary infections (and deaths) as compared to countries with smaller households.

**Figure S2.**
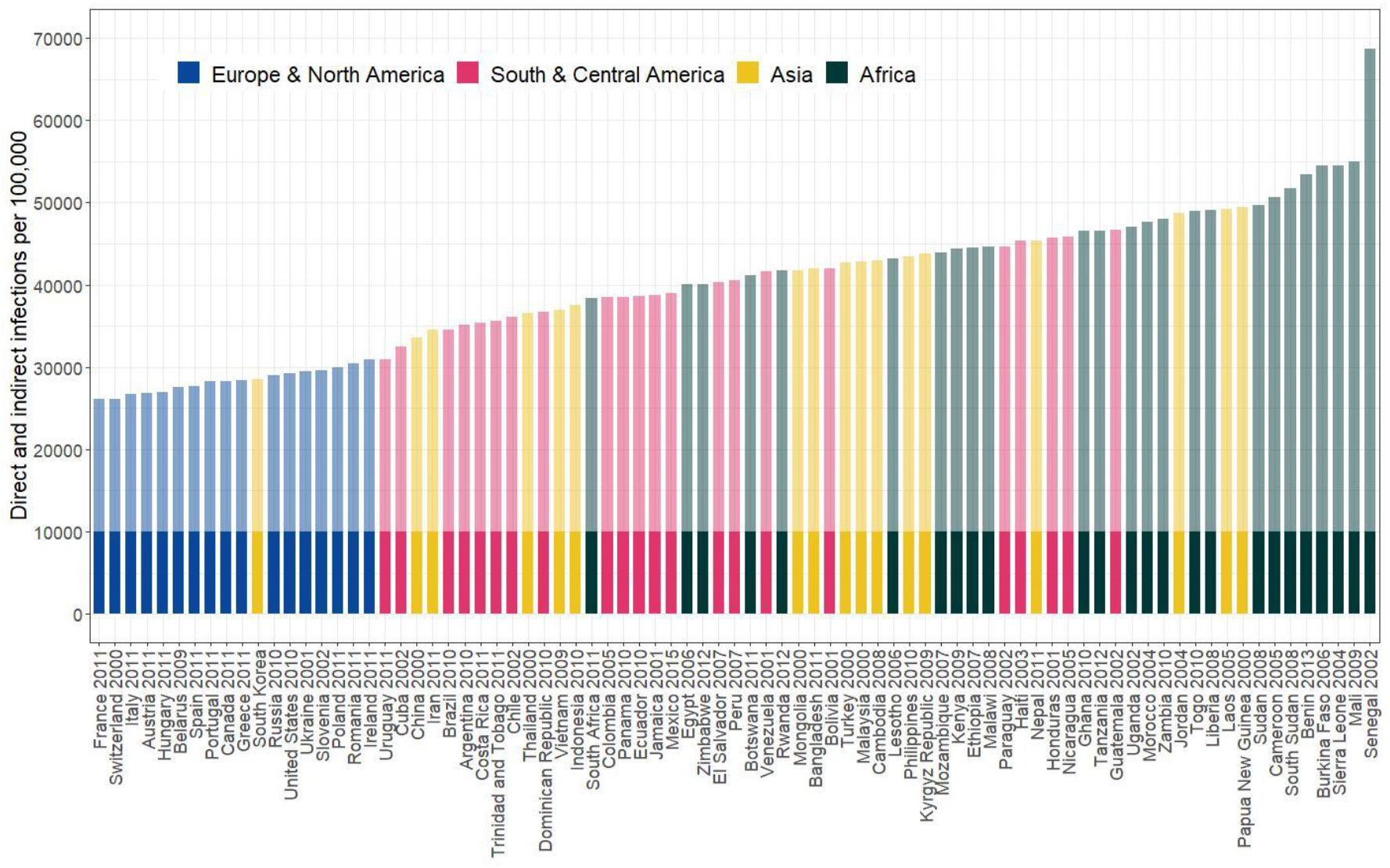
Estimates of primary and secondary infections instead of deaths. Figure S2 displays the simulated infection rates per 100,000 population in our main analysis. As 10,000 individuals are infected in all countries, primary rates do not vary. The secondary rates vary and the sum of direct and indirect rates divided by 10,000 reflects the average household size in each country. Note that a random primary infection of 10% of the population leads in several countries to the majority of the population being infected after secondary infection.

**Figure S3.**
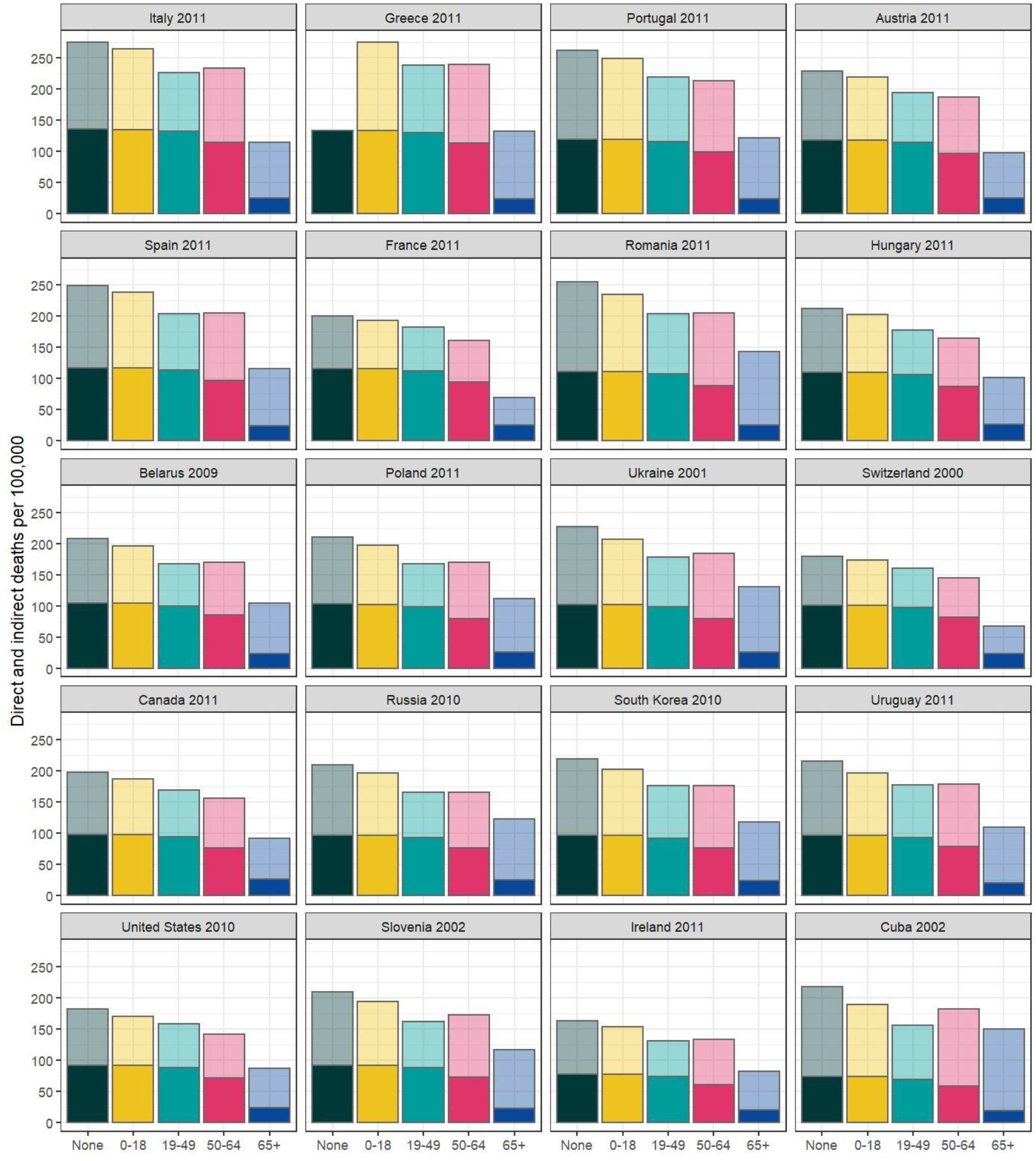
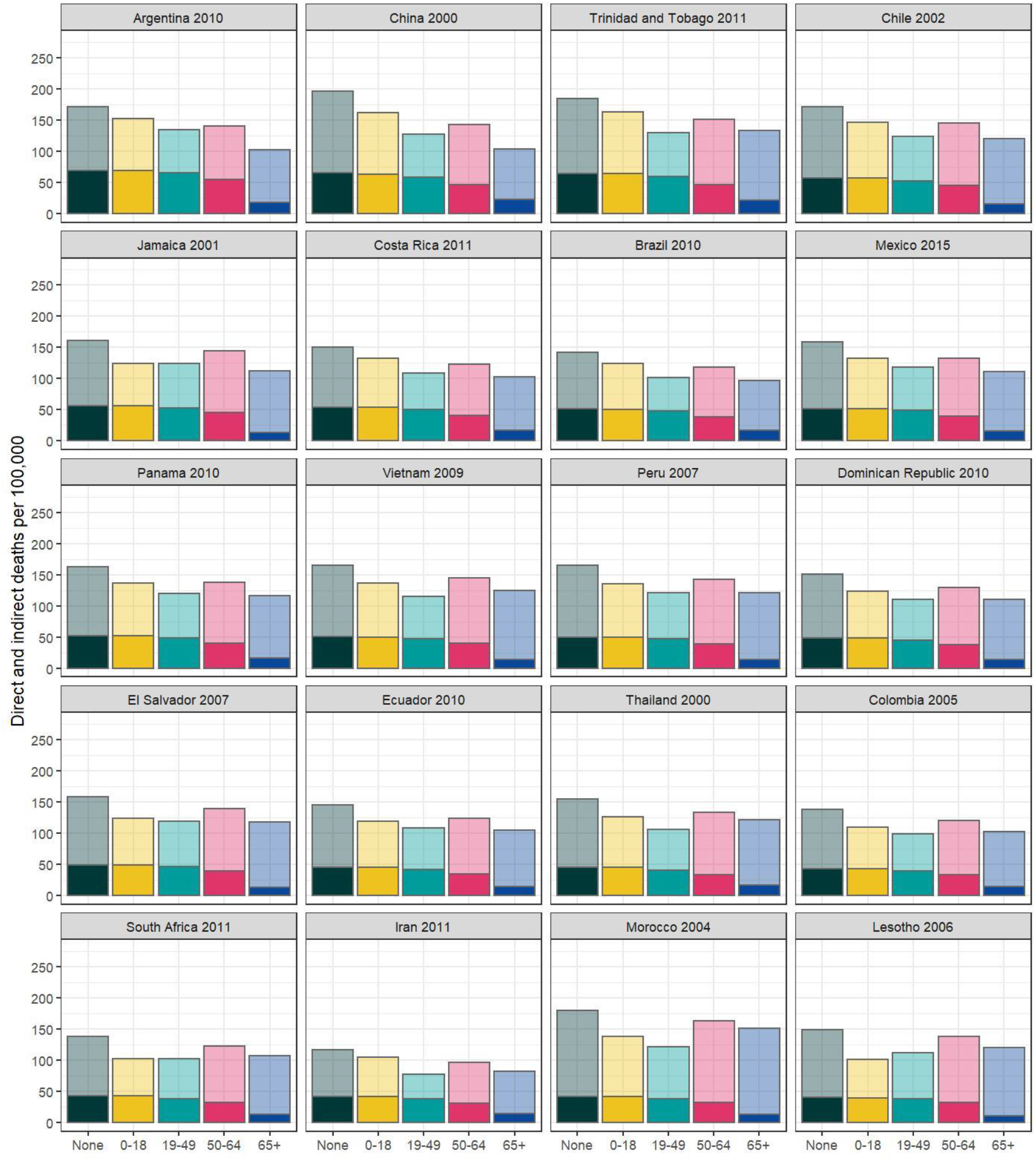
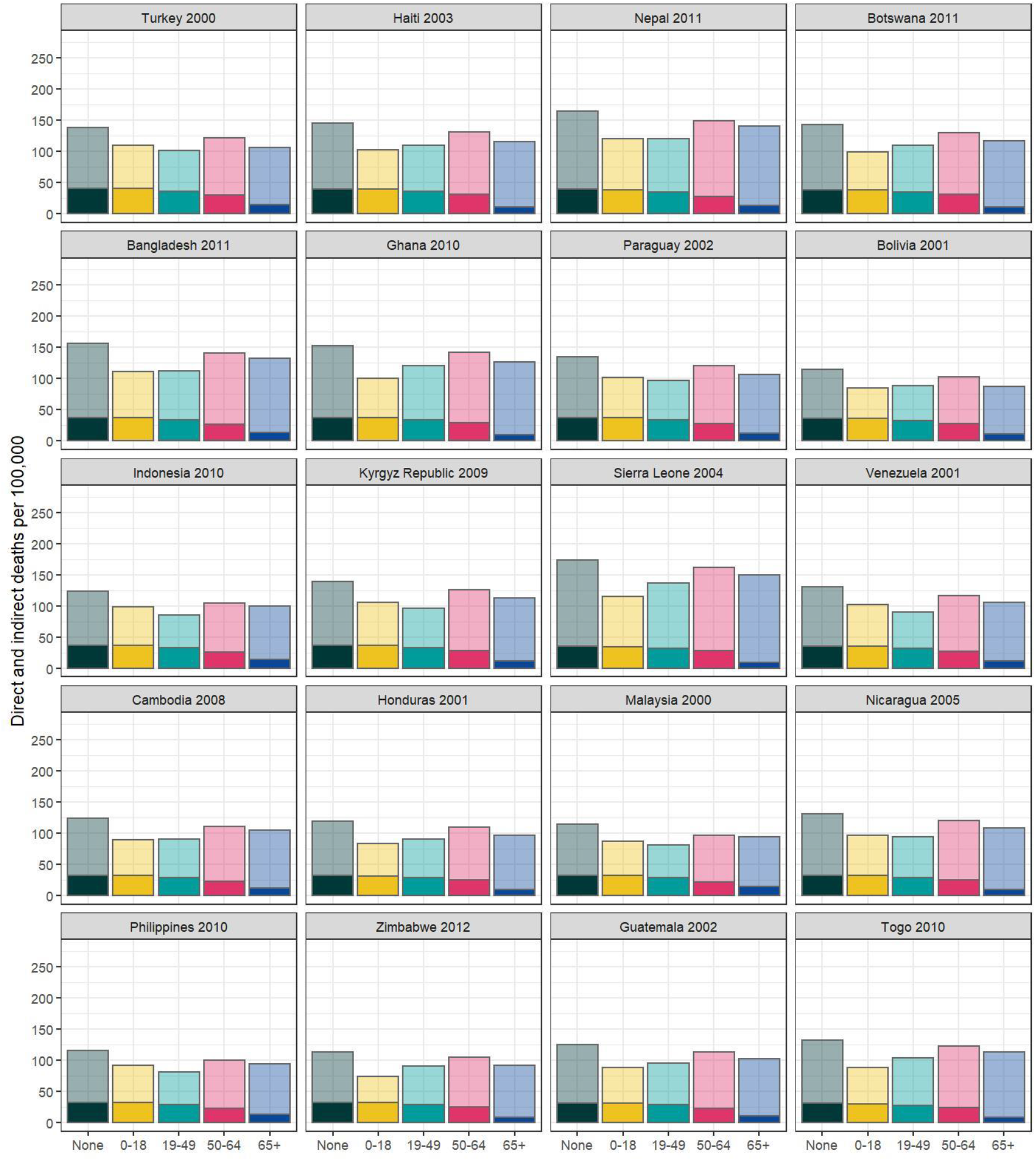
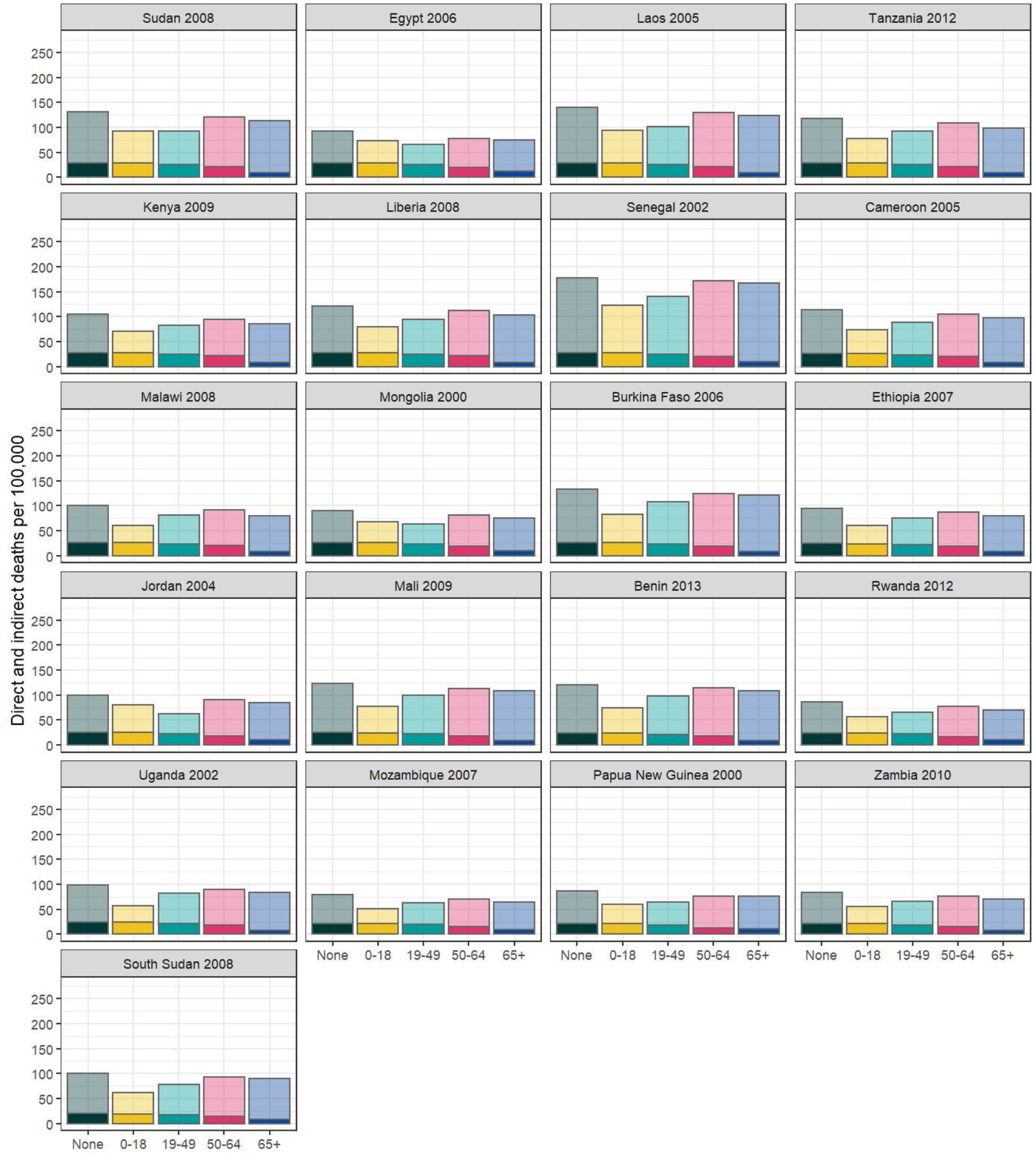
Estimated number of direct (dark) and indirect (light) deaths per 100,000 individuals if primary infections of specific age groups are avoided. Analysis of deaths by household type

**Figure S4.**
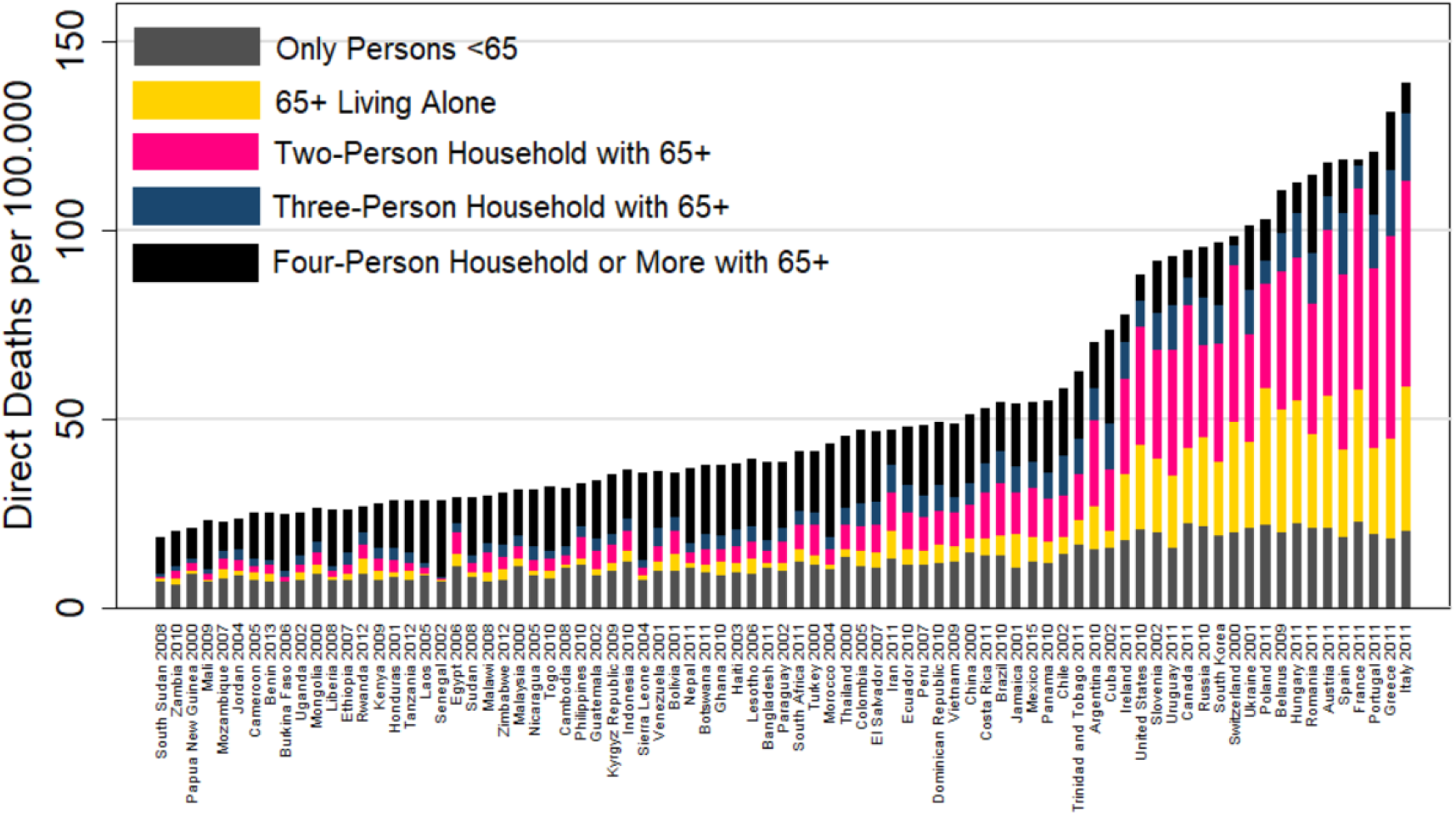
Direct deaths by 100, 000; by living arrangements.

**Figure S5.**
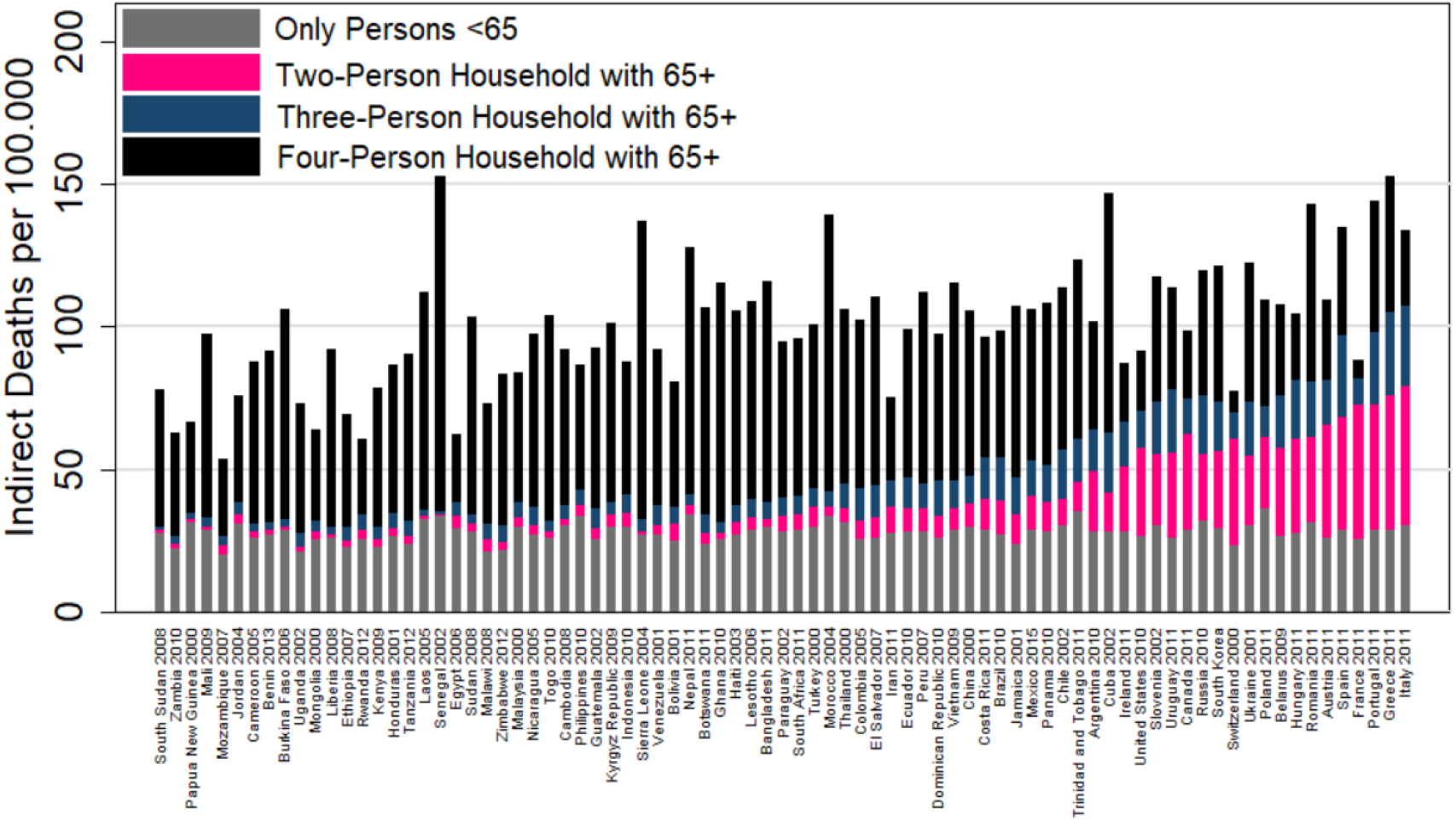
Indirect deaths by 100, 000; by living arrangements. As the number of indirect deaths depends on whom the elderly live with, we provide more insight into the household structures of persons who are simulated to die due to infection with the virus. Figure S4 shows the household structure of the persons who were simulated to die after primary infection (direct deaths). It becomes clear that many of the persons who die due to primary infection are persons aged 65+ who live alone or with another person (probably their partner) in European and North American countries. In Africa and Asia, the majority of direct deaths occur within households where persons above 65+ live with three other persons or more. Figure S5 further completes this picture by showing the household structure of persons who are estimated to die due to secondary infection. These numbers underline that in Europe and North America many indirect deaths occur in households where persons aged 65+ live with one other person (most likely their partner). In France and Switzerland this is the most common household type among those simulated to die due to secondary infection. Not only does this appear to reduce the number of estimated indirect deaths, it might also make confining the elderly in their households a more effective strategy to further reduce indirect deaths. In contrast, in Asia and Africa the majority of indirect deaths occur in households where a person over 65 lives in a household with four or more members. As indirect deaths comprise the lion’s share of estimated deaths in these countries, reducing mortality in Africa and Asia will depend on how effectively secondary infections within large households can be prevented.

### Analysis adjusting for sex

Initial reports indicated that men die more often after being infected with Covid-19 than women. Fatality/infection ratios were not available to us by age for males and females separately. Because men are underrepresented in older age groups, it is not straightforward to recalculate age and sex specific fatality ratios from the separate pieces of information. We therefore decided to only adjust for age in the main analysis and to check how robust these results were as follows. The Chinese Center for Disease Control and Prevention reported that fatalities for men were 2.8% of infections and for women 1.7% (http://weekly.chinacdc.cn/en/article/id/e53946e2-c6c4-41e9-9a9b-fea8db1a8f51). In robustness checks, we therefore adjusted age-specific fatality rates by sex such that the male death rate was 1.24 times the rate for the population as a whole and the female death rates was 0.76 times the population rate.

Figure S6 shows little change in terms of ranking of countries. There is a slight reduction in deaths because of the under-representation of men in older age groups, which makes the above calculation adjust overall fatality rates downward.

**Figure S6.**
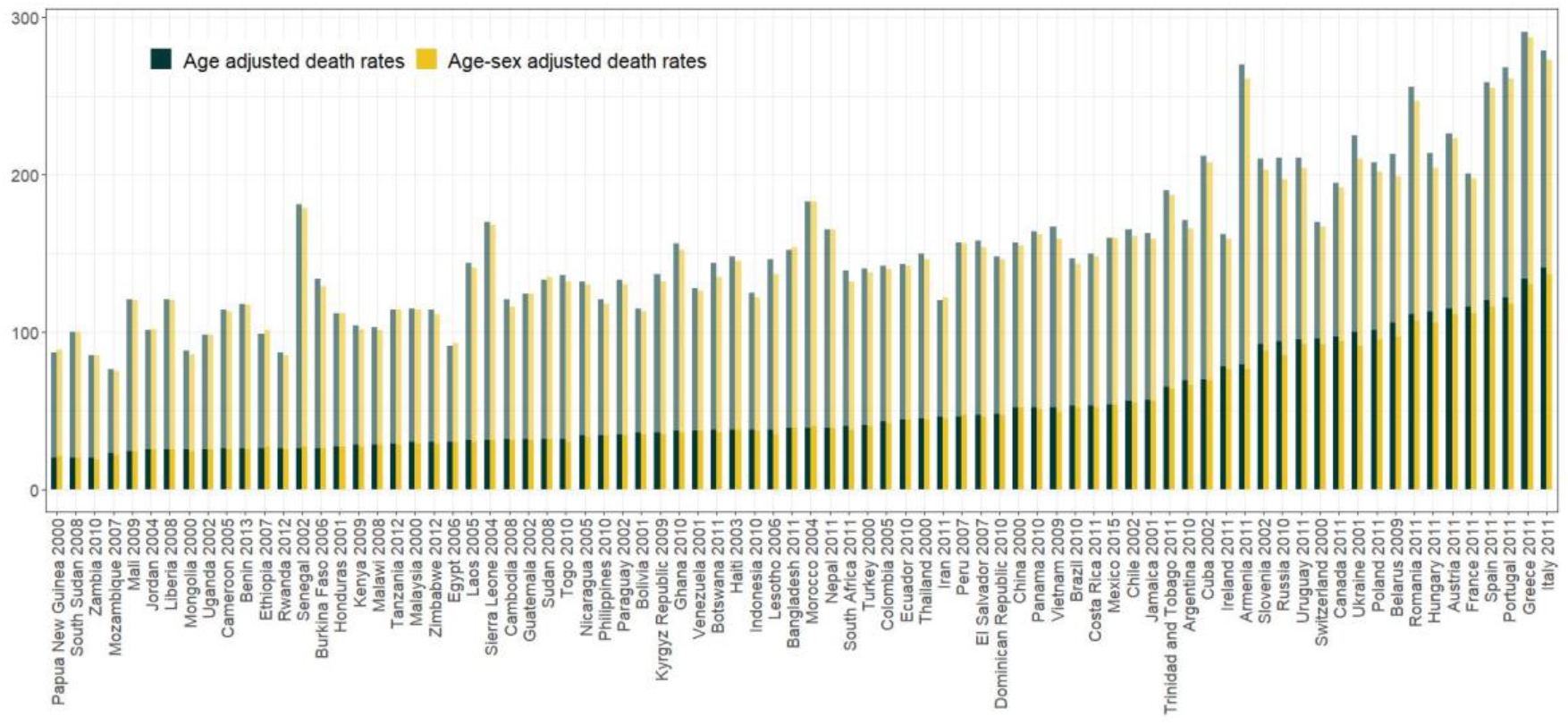
Comparison of direct and indirect death rates per 100,000 using age-adjusted and age-sex-adjusted fatality rates

### Analysis using alternative infection fatality ratios

In our main analysis, we use the death rates as reported by Verity and colleagues.^1^

Ages 0.9: 0.002%; Ages 10–19: 0.006%; Ages 20–29: 0.03%; Ages 30–39: 0.08%; Ages 40–49:

0.15%; Ages 50–59: 0.6%; Ages 60–69: 2.2%; Ages 70–79: 5.1%; Ages 80+: 9.3%.

In robustness checks, we replicated the main analysis using updated Infection Fatality Ratios as reported in Verity and Colleagues^2^, which are slightly lower for older ages. These updated fatality ratios are:

Ages 0.9: 0.00161%; Ages 10–19: 0.00695%; Ages 20–29: 0.0309%; Ages 30–39: 0.0844%; Ages

40–49: 0.161%; Ages 50–59: 0.595%; Ages 60–69: 1.9%; Ages 70–79: 4.3%; Ages 80+: 7.8%.

Figure S7 and S8 compare results using these different fatality ratios. Estimates of the total number of deaths per 100.000 population are reduced to some extent, but differences among countries change very little.

**Figure S7.**
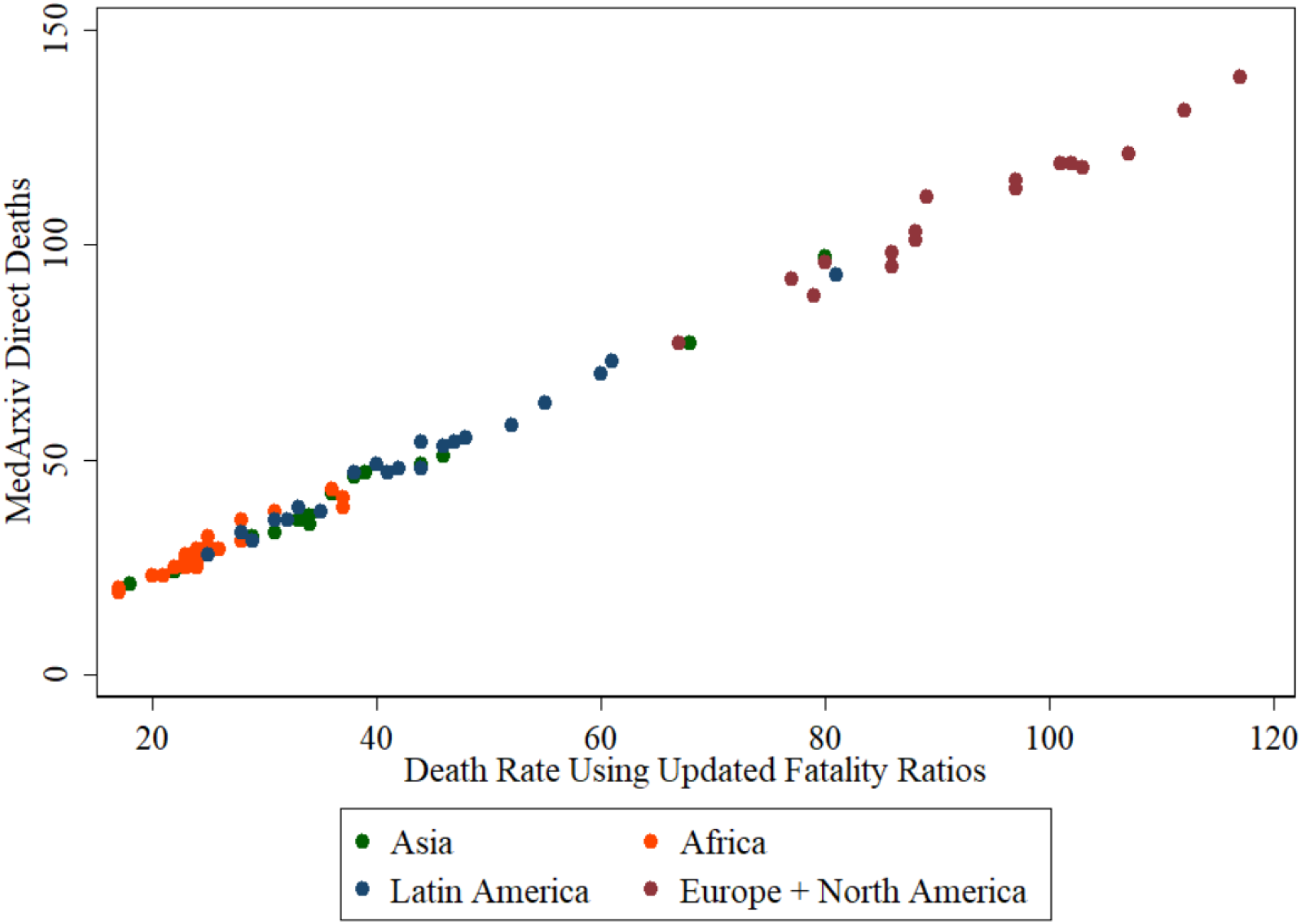
Comparison of direct death estimates using original and updated fatality rates

**Figure S8.**
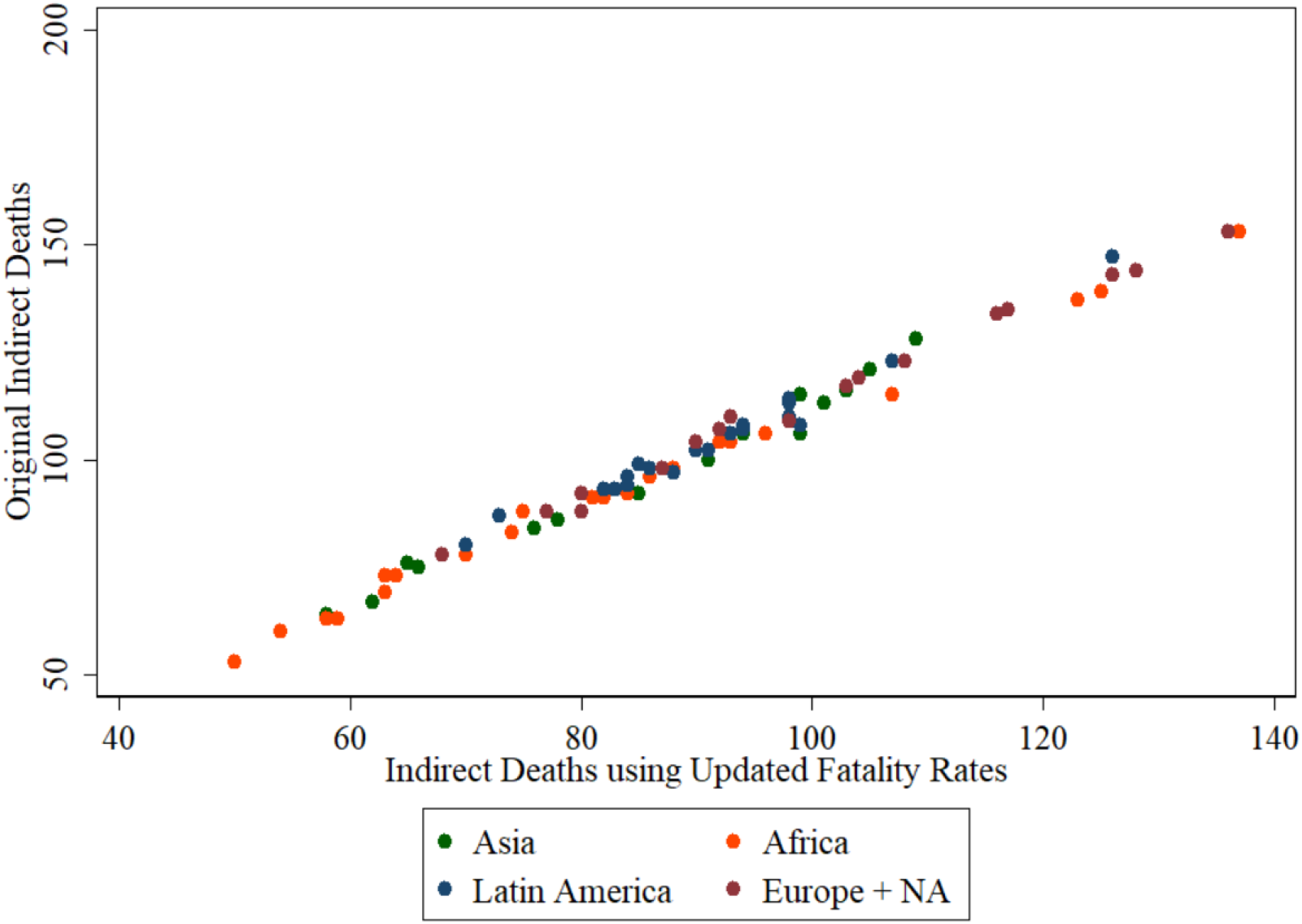
Comparison of indirect death estimates using original and updated fatality rates

### Group Quarters

Only a subset of datasets included individuals living in group quarters. For our purposes, this is especially relevant in the case of residences for the elderly. Therefore, we compared changes in estimates when including them in the subset of countries for which data was available. In some datasets large households are split into various 1-person households. These cases are excluded from both sets of estimates.

Figure S9 shows how including group quarters leads to small upward adjustments in countries like Italy and Switzerland where many old people live in group quarters. In other counties small downward adjustments are observed as many young people live in group quarters (e.g. Jamaica).

**Figure S9.**
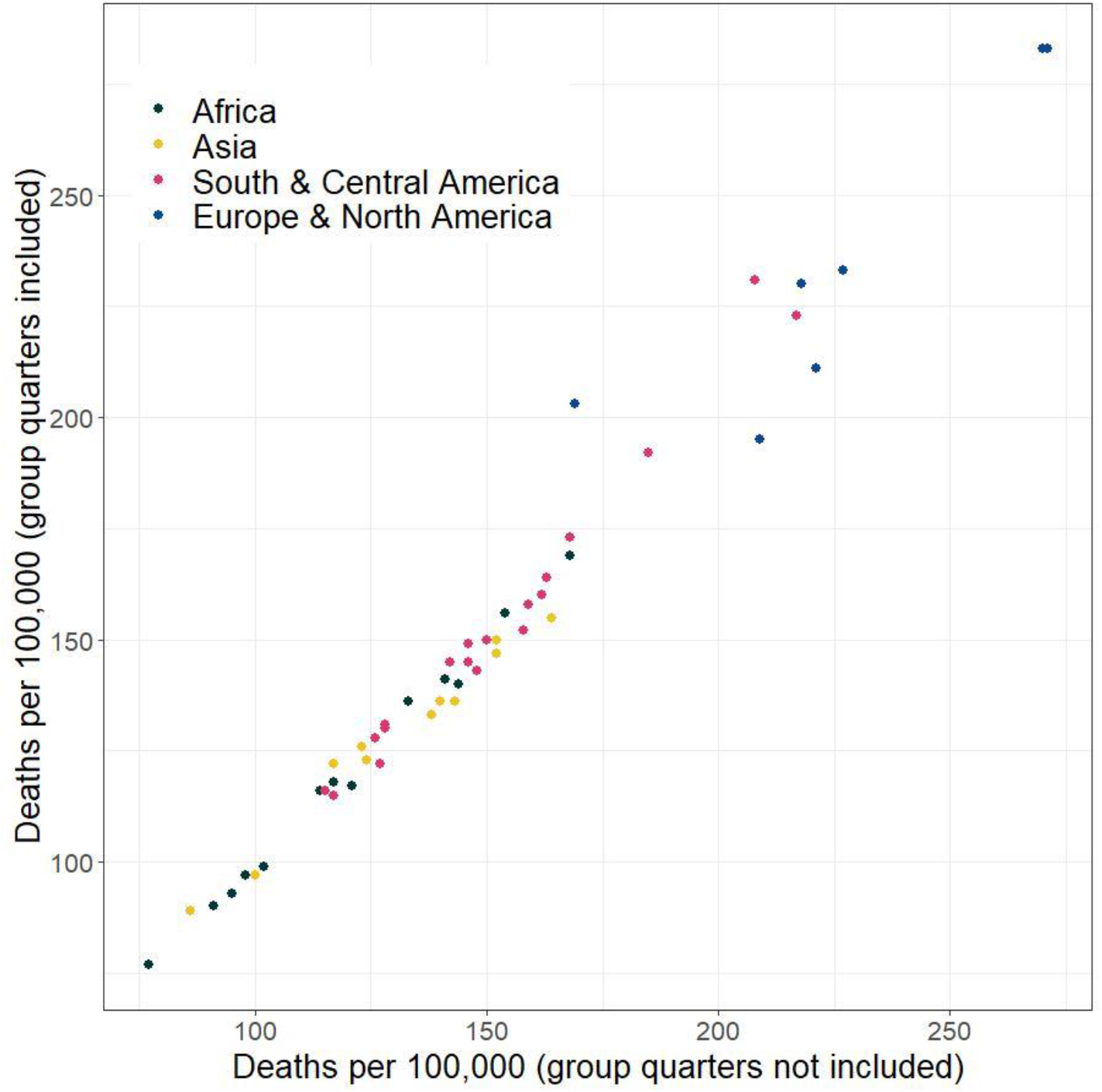
Comparison of total death estimates including and excluding group quarters

### Repeated Random Draws to Simulate Primary Infections

The analysis relies on simulating the random infection of 10,000 individuals out of 100,000 in each sample. This implies that running the analysis several times can lead to slightly different estimates. To investigate the sensitivity of estimates to this issue, we re-ran the analysis 1000 times for Spain. Figure S10 and S11 display the distribution of estimated direct and indirect deaths per 100.000 persons resulting from these 1000 repetitions. It can be observed that the estimated number of deaths varies across random draws, but these changes are relatively small. Even though this might affect the specific ranking of countries that are very similar in terms of the total simulated deaths, no major changes of countries in the death distribution is likely to arise from this issue.

**Figure S10.**
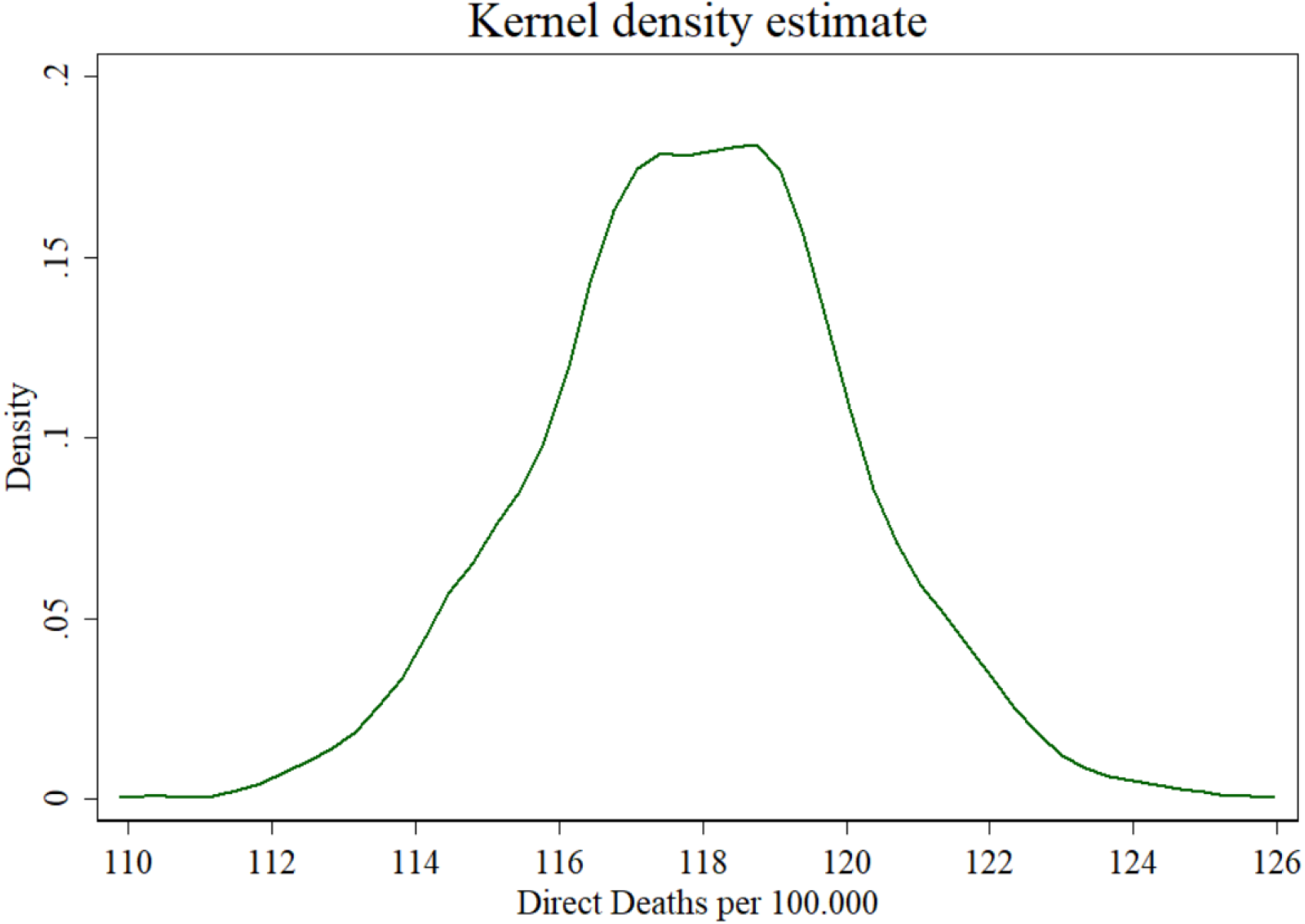
Estimated direct deaths per 100.000 persons resulting from 1000 repetitions of analysis for Spain

**Figure S11.**
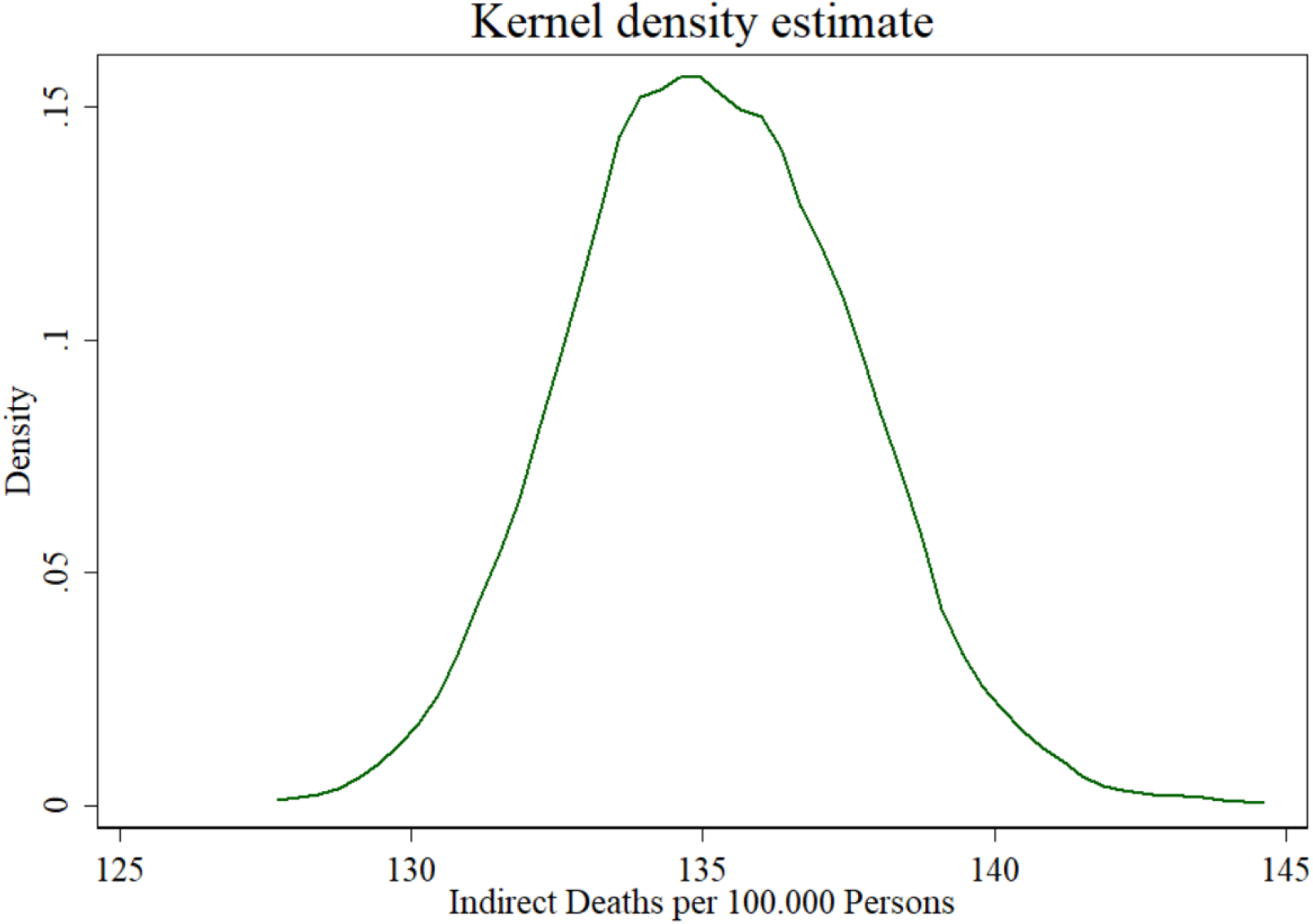
Estimated indirect deaths per 100.000 persons resulting from 1000 repetitions of analysis for Spain

### Adjustment for Italy

The census data of Italy does not distinguish ages above 75. We therefore used ISTAT data from Multiscopo for the year 2009 to divide those above age 75 into two groups: those aged 75–79 and those aged 80 or more. In the 2009 Multiscopo data, 55.9% of those 75+ are aged 80 or more. The website of the Italian Statistical Office ISTAT (http://dati.istat.it/#) provides data from 2012 onward, which indicated a similar percentage of 59.4% of those 75 or older being 80+ for 2012. We took the more conservative estimate (in terms of simulated deaths it would produce) and randomly assigned 55.9% persons to be above 80. A direct comparison with assigning 59.4% of those 75 or older to be 80+ showed very similar estimates:

55.9% 80+: 141 direct deaths and 134 indirect deaths

59.4% 80+: 142 direct deaths and 136 indirect deaths

### Adjustment of China 2000 data to 2010 age structure

IPUMS only provides Chinese Census data for the year 2000. Given the importance of China as the first country where the virus spread to a large part of the population, we decided to adjust estimates to 2010 age structures. This makes the estimates of our main analysis more comparable to the data sources used for other countries which mostly refer to the years 2007-2011. All robustness checks use data from China 2000.

To adjust for changes in the age structure we used data on age by year from the 2010 census as available at (accessed 31/03/2020):

https://www.census.gov/data-tools/demo/idb/region.php?T=10&RT=0&A=both&Y=2010&C=CH&R=

We first ran the analysis using the 2000 census and calculated individuals’ risk of direct and indirect death by age. This risk was calculated for each age in years, but grouped for those aged 80–84 and 85 or older to gain more reliable estimates. We subsequently took the age distribution from the 2010 census, and multiplied the age-specific direct and indirect death risks with the absolute population size of each age. Finally, we summed all deaths across age groups and calculated the number of deaths and indirect deaths per 100.000 population. This exercise was repeated for the simulations where specific age groups were excluded from primary infection.

Because the population of China aged between 2000 and 2010, estimated deaths are higher after these adjustments are made:

|  | *China 2000; Unadjusted* |  | *China 2000; Adjusted to 2010* |  |
| --- | --- | --- | --- | --- |
| *Group without primary infections* | *Direct* | *Indirect* | *Direct* | *Indirect* |
| None | 52 | 108 | 65.9 | 130.4 |
| Aged 0-18 | 52 | 83 | 62.8 | 98.7 |
| Aged 19-49 | 48 | 64 | 58.6 | 69.2 |
| Aged 50-64 | 39 | 96 | 46.6 | 96.1 |
| Aged 65+ | 18 | 101 | 22.2 | 81.3 |

## CODE AVAILABILITY

STATA code used for data management and analysis.

*** Register and Download data from https://international.ipums.org/international/

*** For each country, select the most recent sample since 2000.

*** Netherlands and UK are not selected, which do not have data organized according to

households.

*** Fiji, Puerto Rico, and Togo are not selected due to sample size <100000

*** For the US, we did not use the ACS data available in IPUMS international, but the 2010

census available at IPUMS USA

*** Data for South Korea come from KOSTAT

*** In IPUMS select the variables SAMPLE SERIAL HHWT GQ REGIONW GEO1_FR2011 PERNUM

AGE SEX

*** To make the datafile easier to digest for your computer, use the option “Customize Sample

Sizes” and select at least 110.000 individuals per country

**** set a path where to save files

cd “set path here”

use “200323 todos los paises.dta”, clear

*** drop overseas regions from France and subsequently the variable

drop if geo<10

drop geo

*** drop any implausible values on age

drop if age>120

**** drop individuals living in group quarters

**** for a robustness check including those in group quarters only exclude 1-person units

created by splitting large households and unknown: keep if gq<29

keep if gq==10

drop gq

************************************

***** The following code serves to select households until 100,000 individuals are reached

************************************

**** Most datasets are self-weighting according to IPUMS (Can be found in “Microdata sample

characteristics” once clicking on a specific sample

**** For several samples a stratified selection procedure has been used for data collection, or

other methods that require the inclusion of household weights.

**** For these sample, we created x duplicates of each individual case, x being the household

weight. To avoid an enormous dataset, we divided weights by 100 or 10 depending on the

minimum weight (if minimum weight<100 we used 10)

replace hhwt=hhwt/100 if sample==76201001 | sample==704200901 | sample==764200001 |

sample==834201201 | sample==170200501 | sample==124201101 | sample==250201101 |

sample==364201101 | sample==608201001 |

sample==724201101

replace hhwt=hhwt/10 if sample==388200101 | sample==728200801 | sample==729200801

expand hhwt if sample==76201001 | sample==704200901 | sample==764200001 |

sample==834201201 | sample==729200801 | sample==728200801 | sample==388200101 |

sample==170200501 | sample==124201101 | sample==250201101 | sample==364201101 |

sample==608201001 | sample==724201101, g(dupl)

********************** after expanding the data we created unique household numbers for

each duplicate. This all to create a pool of households from which the sample is drawn to reach

100.000 households

sort sample serial pernum

by sample serial pernum: gen dupno=_n

tostring serial, g(hhnr)

tostring dupno, g(d)

gen hhid=hhnr+”d”+d

drop serial hhnr d dupno dupl

rename hhid serial

**********************************************************

*** To randomize the draw we generated a random number between 0 and 1 for each first

person in the household.

sort sample serial

by sample serial: gen order=_n

sort sample order

by sample order: gen randomize=uniform() if order==1

*** Subsequently we sort the sample by this random number and assign a random household

rank to the first person of each household (rand2)

sort sample order randomize

by sample order: gen rand2=_n if order==1

*** Apply the household rank to all persons of the household

sort sample serial

by sample serial: egen rand3=max(rand2)

*** Create a person rank for each sample

sort sample rand3

by sample: gen ccc=_n

**** generate a marker for the lowest person rank within each household

sort sample serial

by sample serial: egen cut=min(ccc)

*** drop all households with a minimum person rank above 100.000. This will generate samples

of 100.000 persons are a few persons more

drop if cut>100000

*** drop unnecessary variables

drop order randomize rand2 rand3 ccc cut

*************************************************************

***** Save the selected sample

save “200104 sample_selected.dta”, replace

******

use “200104 sample_selected.dta”, clear

**** add any eventual extra datasets. In our case US 2010 and South Korea 2010

append using “200323 USappend.dta”

append using “SouthKorea.dta”

label define sample_lbl 201007 “United States 2010”, add

label define sample_lbl 1111 “South Korea”, add

****************************

**** For Italy, ages above 75 are grouped together. Our analysis depends death rates that vary

across 10-year groups but

**** above 80 there is no further distinction in death rates. We therefore randomly divided

individuals for Italy across the 70s and 80s groups

**** based on the distribution observed in ISTAT’s Multiscopo 2009: Among individuals aged

75+ 55.9% is aged 80+

sort sample age

by sample age: gen ao=uniform() if sample==380201101 & age==75

replace age=85 if sample==380201101 & age==75 & ao<0.5586

**** insert deathrate by age in code below

******* deathrate from imperial college london https://www.imperial.ac.uk/media/imperial-college/medicine/sph/ide/gida-fellowships/Imperial-College-COVID19-NPI-modelling-16-03-2020.pdf

gen deathrate=0.00002 if age>−1 & age<10

recode deathrate .= 0.00006 if age>9 & age<20

recode deathrate .= 0.0003 if age>19 & age<30

recode deathrate .= 0.0008 if age>29 & age<40

recode deathrate .= 0.0015 if age>39 & age<50

recode deathrate .= 0.0060 if age>49 & age<60

recode deathrate .= 0.022 if age>59 & age<70

recode deathrate .= 0.051 if age>69 & age<80

recode deathrate .= 0.093 if age>79 & age<120

*** In robustness checks we adjusted for sex using a uniform formula:

*** replace deathrate=deathrate*1.24 if sex==1

*** replace deathrate=deathrate*0.76 if sex==2

**********************

*** The following code randomly selects individuals who are infected

******* insert amount of absolute cases infected after local cases. 10000 represents 10%; In

robustness checks we infected 20000.

local cases 10000

foreach x of local cases{

sort sample

by sample: gen random=runiform()

sort sample random

by sample: gen order=_n

gen infected=1 if order<=‘x’

recode infected .= 0

}

**********************

**** generate indicator of households with at least one infected individual

sort sample serial

by sample serial: egen infhh=max(infected)

*** indicator of the amount of infected individuals in a given household

sort sample serial

by sample serial: egen amtinfhh=total(infected)

*** random number to select share of uninfected who get infected by household members, this

code is only necessary if one assumes less than 100% of hh members get infected

sort sample serial infected

by sample serial infected: gen infran=runiform() if infected==0

**** In our analysis, we assumed that all household members get infected (or to show the

maximum impact household transmission can have)

**** If you would like to use a different within household transmission rate, insert this after

“local hhrate 0” as a percentage out of 100, but keep the 0 (e.g. “local rate 0 50”)

**** This percentage indicates the probability that an infected individual infects an uninfected

household member

**** In other words, the probability of an uninfected person becoming infected increases with

the number of primary infections in the household

**** The numbers that emerge from hhrate 0 assume no secondary infections and indicate

deaths due to primary infections only

local hhrate 0 100

**** chance of being infected depending on hh members infected

foreach y of local hhrate{

gen infchance‘y’= 1–((1–(‘y’/100))^amtinfhh)

**** generate an indicator of all persons infected after primary and secondary infections

gen finalinfect‘y’=1 if infran<= infchance‘y’ & infected==0 & infchance‘y’! =.

replace finalinfect‘y’ = 1 if infected==1

*** generate the total number of primary plus secondary infections per 100.000 in each country

by sample: egen totinf‘y’=total(finalinfect‘y’)

********* calculate the amount of deaths per 100.000 in each country

*** individual chance of dying by multiplying with 1 the death rates if a person is infected

gen deathchance‘y’=deathrate*finalinfect‘y’

**** sum all individual death chances within each sample

sort sample

by sample: egen amtdeath‘y’=total(deathchance‘y’)

**** generate integers for presentation purposes

gen amtdeathint‘y’=round(amtdeath‘y’)

}

**** generate estimate of direct and indirect deaths per 100.000

gen direct=amtdeathint0

gen indirect=amtdeathint100-amtdeathint0

*** indicator for total deaths per 100.000

gen totdeath=amtdeathint100

*** drop unnecessary variables

drop random order infhh amtinfhh infran infchance0 totinf0 amtdeath0 amtdeathint0

infchance100 totinf100 amtdeath100 amtdeathint100

**** numbers for Figure 1 in Table. Graph bar to see all is ok.

graph bar direct indirect, stack over(sample, sort(direct))

table sample, c(mean direct mean indirect mean regionw) format(%9.0g)

******

save “200401 comparative.dta”, replace

**** alternative way of creating data for Figure 1 using collapse which maintains region labels

use “200401 comparative.dta”, clear

collapse (mean) direct indirect regionw, by(sample)

label values regionw regionw_lbl

recode regionw .= 23 if sample==201007

recode regionw .= 32 if sample==1111

********* Data for Supplementary Figure on Infections

use “200401 comparative.dta”, clear

collapse (sum) finalinfect0 finalinfect100 regionw, by(sample)

gen indirectinf=finalinfect100-finalinfect0

drop finalinfect100

label values regionw regionw_lbl

recode regionw 0=23 if sample==201007

recode regionw 0=32 if sample==1111

******************* Additional Code for Supplementary Analysis on Household Types Among

65+ Fatalities

use “200401 comparative.dta”, clear

*** generate indicators splitting the sample into <65 and 65+

gen old=1 if age>64 & age!=.

recode old .= 0 if age<65

*** generate variable with number of 65+ in each household

sort sample serial

by sample serial: egen nold=total(old)

*** generate variable with total number of household members

sort sample serial

by sample serial: gen hhsize=_N

**** generate household classification

gen hhtype=0 if nold==0

recode hhtype .= 1 if nold>0 & hhsize==1 & nold!=.

recode hhtype .=2 if nold>0 & hhsize==2 & nold!=.

recode hhtype .=3 if nold>0 & hhsize==3 & nold!=.

recode hhtype .=4 if nold>0 & hhsize>3 & nold!=. & hhsize!=.

***** generate variables that only contain the probability of a person infected in the random

draw (deathchance0)

*** or after secondary infections (deathchance100) dying if a person lives in a given household

type.

gen dh0= deathchance0 if hhtype==0

gen ih0=deathchance100 if hhtype==0

gen dh1= deathchance0 if hhtype==1

gen ih1=deathchance100 if hhtype==1

gen dh2= deathchance0 if hhtype==2

gen ih2=deathchance100 if hhtype==2

gen dh3= deathchance0 if hhtype==3

gen ih3=deathchance100 if hhtype==3

gen dh4= deathchance0 if hhtype==4

gen ih4=deathchance100 if hhtype==4

**** drop individuals below 65

**** create a dataset that indicates direct and total deaths among 65+ by household type

collapse (sum) dh0 ih0 dh1 ih1 dh2 ih2 dh3 ih3 dh4 ih4 regionw direct, by(sample)

*** generate indicator for indirect deaths

gen i0=ih0-dh0

gen i1=ih1-dh1

gen i2=ih2-dh2

gen i3=ih3-dh3

gen i4=ih4-dh4

***** crappy graphs to check things worked, sorted by direct deaths.

graph bar dh0 dh1 dh2 dh3 dh4 if sample!=51201101, stack over(sample, sort(direct))

graph bar i0 i1 i2 i3 i4 if sample!=51201101, stack over(sample, sort(direct))

******************************************************************************

****

************* ANALYSIS OF AVOIDING PRIMARY INFECTIONS OF AGE GROUPS

**************

******************************************************************************

****

use “200401 comparative.dta”, clear

**** divide sample into age groups

gen agegroups=0 if age<19

recode agegroups .=1 if age>18 & age<50

recode agegroups .=2 if age>49 & age<65

recode agegroups .=3 if age>64 & age<120

label define ag 0 “0–18 years” 1 “19–49 years” 2 “50–64 years” 3 “6+ years”, replace

label values agegroups ag

**** rename the original infected variable to set original 10.000 infected for each

counterfactual

rename infected infected2

**** drop other previous infection and death variables

drop finalinfect* deathchance*

**** insert percentage of cases in household that eventually become infected after hhrate

local hhrate 0 100

********* if you use alternative age groups, insert their values after “local agegr”

local agegr 0 1 2 3

foreach m of local agegr{

****** recode primary infections among specific age group to 0

gen infected=infected2

recode infected 1=0 if agegr==‘m’

**** simulate household transmission with new number of primary infections, same code as

before

**** generate indicator of households with infected individual

sort sample serial

by sample serial: egen infhh=max(infected)

*** indicator of amount infected within household

sort sample serial

by sample serial: egen amtinfhh=total(infected)

*** random number to select share of uninfected who get infected by household members

sort sample serial infected

by sample serial infected: gen infran=runiform() if infected==0

**** chance of being infected depending on hh members infected

foreach y of local hhrate{

gen infchance‘y’=1–((1-(‘y’/100))^amtinfhh)

**** generate variable of all infected after secondary infections

gen finalinfect‘y’ = 1 if infran<infchance‘y’ & infected==0 & infchance‘y’!=. & infran!=.

replace finalinfect‘y’=1 if infected==1

**** total infections in sample

sort sample

by sample: egen totinf‘y’=total(finalinfect‘y’)

********* amount of deaths in sample

gen deathchance‘y’=deathrate*finalinfect‘y’

sort sample

by sample: egen amtdeath‘y’ = total(deathchance‘y’)

gen amtdeathint‘y’=round(amtdeath‘y’)

}

*** generate death rates for each age counterfactual

gen direct‘m’=amtdeathint0

gen totdeath‘m’=amtdeathint100

gen indirect‘m’=amtdeathint100-amtdeathint0

drop infected infhh amtinfhh infran infchance0 finalinfect0 totinf0 deathchance0 amtdeath0

amtdeathint0 infchance100 finalinfect100 totinf100 deathchance100 amtdeath100

amtdeathint100

}

******************************************************************************

************************************************

***** The following code prepares the dataset to create a long-format containing all

counterfactual estimates for each country

***** this code should be adjusted if more age categories are used

sort sample

by sample: gen order=_n if _n<6

**** rates for each group are added to each of the rows 2–5 per country; original results are

kept in row 1

replace direct=direct0 if order==2

replace indirect=indirect0 if order==2

replace direct=direct1 if order==3

replace indirect=indirect1 if order==3

replace direct=direct2 if order==4

replace indirect=indirect2 if order==4

replace direct=direct3 if order==5

replace indirect=indirect3 if order==5

label define ord 1 “All” 2 “Aged 0–18” 3 “Aged 19–49” 4 “Aged 50–64” 5 “Aged 65+” 6 “Aged 65+”,

replace

label values order ord

***** keep only those rows with information

keep if order<6

******************************************************************************

***************************************************

*** some examples of results by age

graph bar direct indirect if sample==380201101 | sample==201007, stack over(order)

over(sample, sort(totdeath))

save “200401 age.dta”, replace

***** create a dataset with results by age for all countries

use “200401 age.dta”, clear

replace regionw=23 if sample==201007

replace regionw=32 if sample==1111

browse sample order direct indirect regionw

keep sample order direct indirect regionw

## Footnotes

1 Verity, R., Okell, L.C., Dorigatti, I., … Ferguson, N.M. (2020a) Estimates of the severity of COVID-19 disease. MedRxiv: https://doi.org/10.1101/2020.03.09.20033357

2 Verity, R., Okell, L.C., Dorigatti, I., … Ferguson, N.M. (2020b) Estimates of the severity of coronavirus disease 2019: a model-based analysis. The Lancet: Infectuous Diseases. https://doi.org/10.1016/S1473-3099(20)30243-7

